# Impact of body mass index on functional recovery after total hip arthroplasty: A prospective study

**DOI:** 10.1101/2025.08.01.25332609

**Authors:** Musashi Ima, Tamon Kabata, Daisuke Inoue, Yu Yanagi, Takahiro Iyobe, Naoya Fujimaru, Satoru Demura

**Author notes:** Corresponding author (TK).

## Abstract

**Introduction:** This study aimed to examine the effects of body mass index (BMI) on functional recovery after total hip arthroplasty (THA).

**Materials and methods:** This prospective case-control study included 269 patients who underwent primary THA. Participants were categorized into normal weight, overweight, and obese groups based on the World Health Organization criteria. Recovery outcomes were assessed across BMI groups using gait measurements and the Japanese Orthopaedic Association (JOA) hip scores.

**Results:** Patients with obesity exhibited slower recovery in walking speed and stride length at 6 months and 1 year than did their less-obese counterparts. At 1 year post-surgery, the JOA hip scores showed no significant differences among the BMI groups, thereby indicating similar satisfaction levels despite initial functional recovery differences.

**Conclusions:** Although patients with obesity faced early recovery challenges, particularly regarding gait, satisfaction with THA outcomes was comparable across all BMI groups at 1 year. These findings highlight the need for personalized management and rehabilitation strategies for optimizing THA outcomes for patients with obesity.

## Introduction

Total hip arthroplasty (THA) is a highly effective treatment for restoring hip joint function in patients with hip osteoarthritis (OA) [1]. The number of THA procedures performed annually is increasing and is expected to rise further [2]. OA may result from congenital factors, such as developmental dysplasia of the hip and femoroacetabular impingement, and acquired factors, including heavy lifting, obesity, and sports activities [3]. Overweight and obesity are known risk factors for OA, and as their prevalence rises, so does the incidence of secondary OA of the hip [4,5]. THA in patients with obesity presents challenges, including increased risks of dislocation, deep infection, bleeding, deep vein thrombosis, and the need for revision surgery [6–9]. Moreover, patients with obesity tended to have longer surgical times and hospital stays [10]. From a biomechanical perspective, obesity is associated with reduced hip range of motion [11] and decreased walking speed [12].

However, the effect of obesity on the functional recovery of patients with OA is poorly understood. Some studies have shown that patients with obesity can achieve functional scores similar to those of patients with no obesity [13], whereas others have reported poorer outcomes [14]. Shibuya et al. reported that the maximum walking speed measured over a 10-meter walk as fast as safely possible is an indicator of post-THA functional recovery [15], and Ohmori et al. reported that a comfortable walking speed of 1.34 m/s or more 1 year post-THA indicates favorable long-term functional recovery [16]. To date, no studies have investigated the effect of body mass index (BMI) on walking recovery after THA using both clinical evaluation and biomechanics.

This study explored the functional recovery differences at 6 months and 1 year post-THA among healthy, overweight, and obese groups based on preoperative status.

## Materials and methods

This prospective case-control study was approved by the Ethics Committee of Kanazawa University (Approval Number: [3669]). All procedures were conducted in accordance with the ethical standards set forth in the 1964 Declaration of Helsinki and its later amendments or comparable ethical standards. Written informed consent was obtained from all patients before participation. Between 11 December 2015, and 7 December 2022, 320 hips underwent primary THA for OA at our institution. Patients with osteonecrosis of the femoral head, inflammatory hip disease, severe hip deformities such as (Crowe types 3 and 4), and bilateral THAs were excluded, resulting in 269 hips included in the study.

All patient information was extracted from electronic medical records, and the patients were classified into three groups according to World Health Organization criteria. Radiographic evaluations confirmed the absence of significant osteoarthritic changes in adjacent joints, including the ipsilateral knee, ankle, and lumbar spine, thereby minimizing the confounding effects of adjacent joint pathologies on gait function. A BMI of <25 kg/m^2^ was defined as normal weight, a BMI of 25–29.9 kg/m^2^ as overweight, and a BMI ≥30 kg/m^2^ as obese.

Preoperative and 1-week postoperative computed tomography (CT) scans were obtained for each patient. The CT images covered the area from the iliac crest to the femoral condyles, with slices cut at a thickness of 1 mm and a pitch of 2.5 mm, generating 160–250 slices per patient based on body size. All CT slices were imported into CT-based templating software (ZedHip, Lexi, Tokyo, Japan) in the Digital Imaging and Communications in Medicine format to create virtual three-dimensional bone models for virtual surgery. This software enabled the accurate measurement of various parameters such as leg length and femoral and acetabular offsets [17,18] (Fig 1).

**Fig 1.**
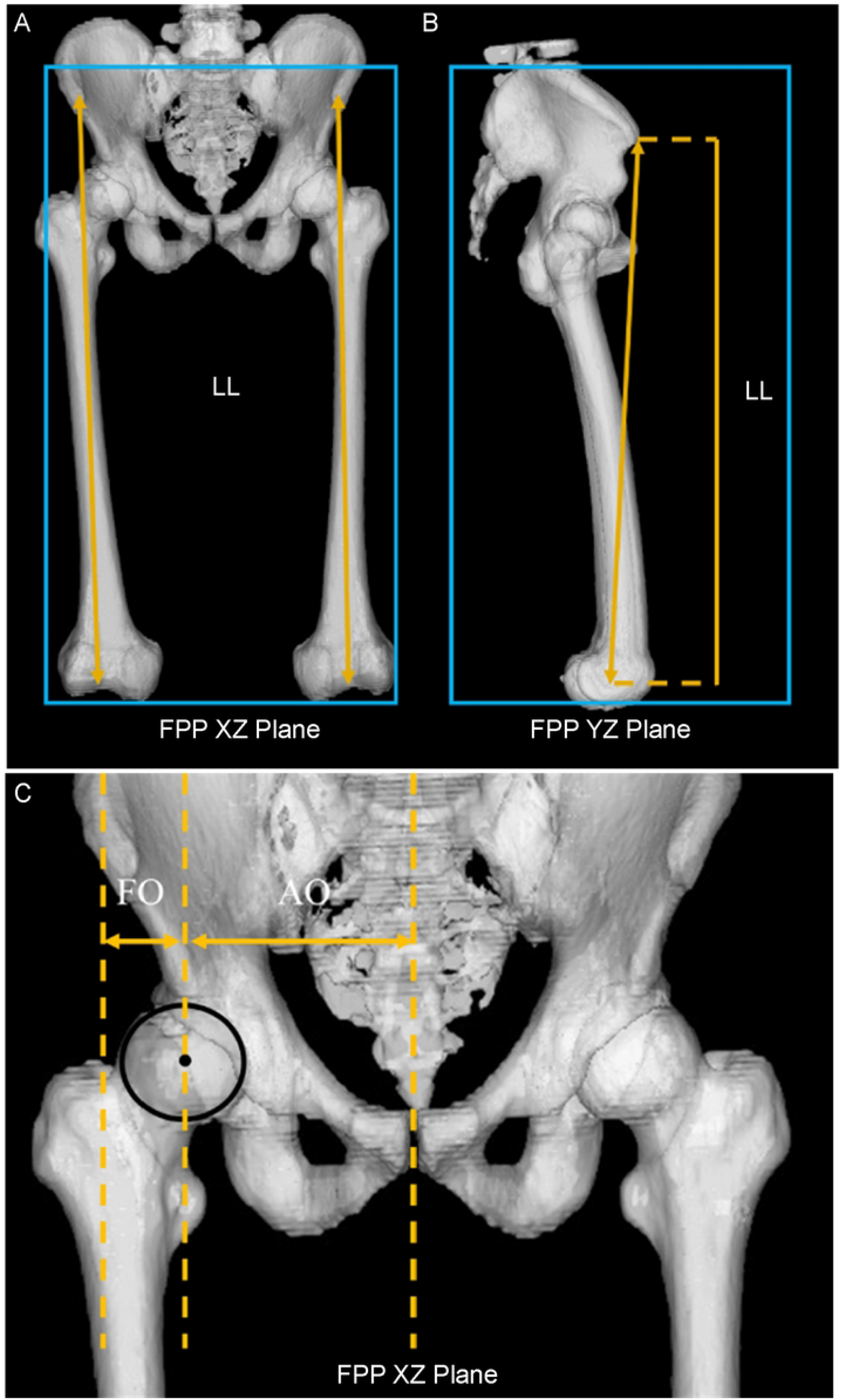
Three-dimensional computed tomography analysis. a and b represent leg length (LL), which is defined as the distance projected onto the frontal plane projection (FPP) from the anterior superior iliac spine to the center of the knee. *c* represents both the femoral offset (FO) and acetabular offset (AO), which are defined as the lengths projected onto the FPP XZ plane in the neutral position. FO is the distance between the center of the femoral head, the center of the cup, and the axis of the femur. AO is the distance between the pubic symphysis and the center of the femoral head or cup. The global offset is the sum of FO and AO.

All surgeries were performed by a single surgeon using an anterolateral approach. The cup was positioned to replicate the center of the contralateral femoral head and medicalized until it contacted the teardrop. The femoral stem was inserted such that its offset increased in proportion to cup medialization, matching the global offset with the contralateral side. The Japanese Orthopaedic Association (JOA) hip score was measured separately using the BMI.

### Assessment: gait measurement

The time required to walk 10 m was measured after a 2-m lead-in (acceleration) phase at a comfortable speed. The time taken to walk 10 m was recorded, and walking speed (m/s) was calculated. Additionally, the stride length (m) was calculated from the number of steps required to cover a 10-m distance [17].

### Statistical analysis

All statistical analyses were performed using the SPSS Statistics version 26 (IBM Corp., Armonk, NY, USA). One-way analysis of variance with repeated measures was used to compare walking speed, stride length, and the JOA hip score between the groups. Sex and American Society of Anesthesiologists status were analyzed using the Chi-square test. Statistical significance was set at p < 0.05.

## Results

### Patient background

Table 1 classifies patients with hip OA into the BMI groups. No significant differences were found between the groups, except for surgical time.

**Table 1.** Patient background.

|  | Healthy group | Overweight group | Obese group | *P*-value |
| --- | --- | --- | --- | --- |
| Age | 64.3±8.8 | 66.1±9.0 | 64.1±8.7 | 0.34 |
| Sex<br>(female/male) | 162/15 | 61/7 | 24/1 | 0.28 |
| BMI | 21.7±2.0 | 27.1±1.4 | 33.9±2.7 | <0.05 |
| Operation time | 154.5±32.5 | 157.5±44.6 | 186.3±21.1 | <0.05 |
| Postoperative day | 17.3±4.6 | 17.2±3.5 | 20.5±3.4 | 0.06 |
| ASA score | 16/150/11 | 0/67/1 | 0/21/3 | 0.11 |
| Blood loss | 219.1±135.8 | 227.7±144.2 | 262.5±123.64 | 0.34 |

BMI: Body mass index; ASA, American Society of Anesthesiologists; operation time, time required for surgery; postoperative day, period of hospitalization; blood loss, intraoperative blood loss

Table 2 presents the preoperative measurements of the leg length discrepancy and offset. No significant differences in leg length discrepancy, acetabular offset, or femoral offset were observed pre- or postoperatively.

**Table 2.** Computed tomography data analysis.

|  | Healthy group | Overweight group | Obese group | *P*-value |
| --- | --- | --- | --- | --- |
| Pre LLD | -8.7±11.0 | -8.7±10.4 | -7.5±9.6 | 0.87 |
| Post LLD | 1.7±6.7 | 0.5±6.2 | 1.0±5.1 | 0.42 |
| Pre acetabular offset | 96.9±9.0 | 96.3±7.1 | 97.4±6.3 | 0.87 |
| Post acetabular offset | 88.5±4.8 | 88.0±4.1 | 88.3±5.2 | 0.42 |
| Pre femoral offset | 27.6±7.2 | 27.8±7.1 | 29.6±7.4 | 0.45 |
| Post femoral offset | 33.1±6.2 | 32.8±4.8 | 32.9±7.5 | 0.95 |

Pre LLD: Preoperative leg length discrepancy
Post LLD: Postoperative leg length discrepancy
Pre acetabular offset: Preoperative acetabular offset
Post acetabular offset: Postoperative acetabular offset
Pre femoral offset: Preoperative femoral offset
Post femoral offset: Postoperative femoral offset

### JOA hip score

Fig 2 shows the progression of the JOA hip score preoperatively, 6 months postoperatively, and 1 year postoperatively. Although no significant differences were found preoperatively, significant differences were observed at 6 months postoperatively between the healthy and obese groups, as well as between the overweight and obese groups. However, no significant differences were found among the three groups at 1 year postoperatively.

**Fig 2.**
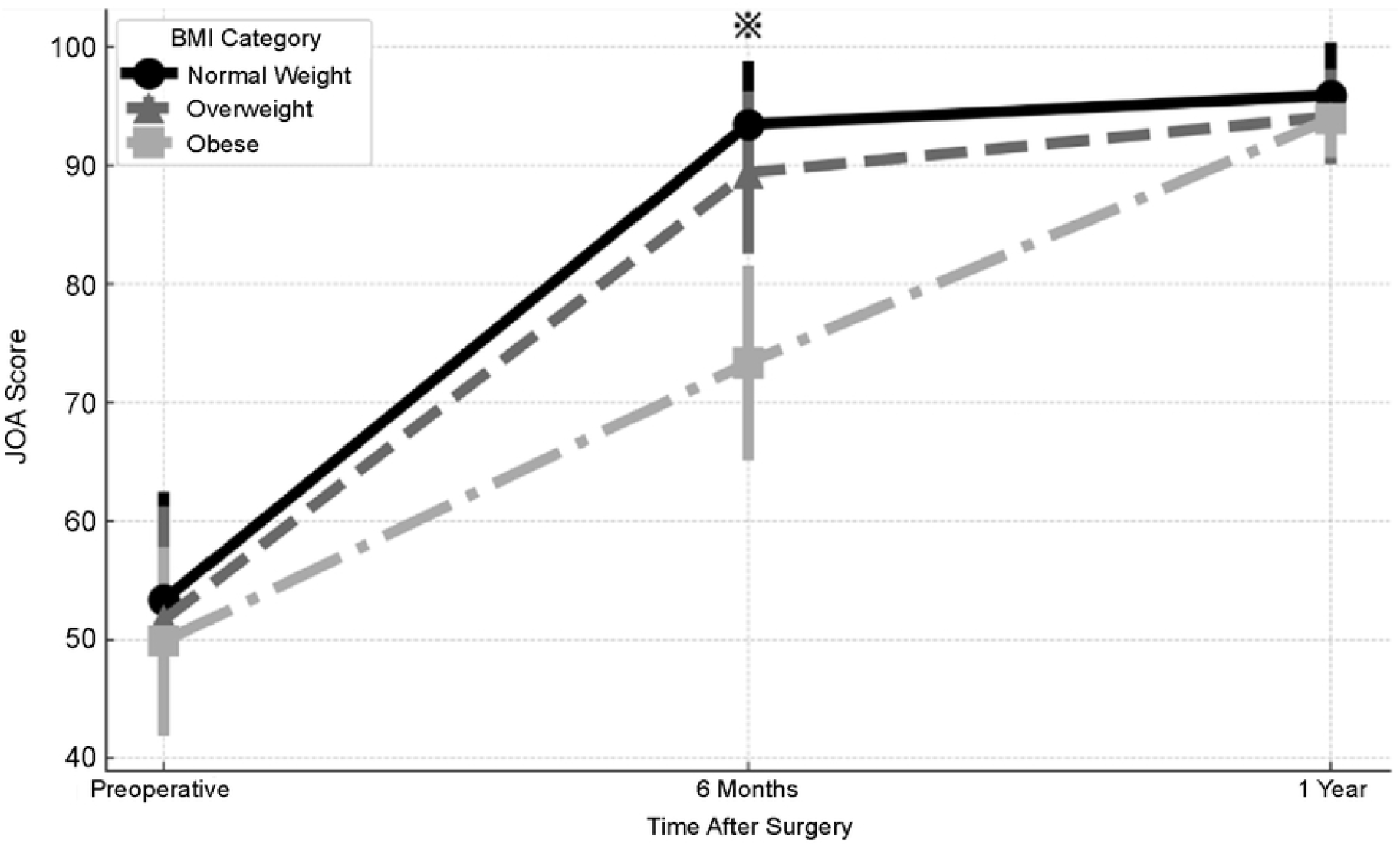
JOA hip score. No significant difference in Japanese Orthopaedic Association (JOA) scores is observed among the three groups at the preoperative and 1 year postoperative time points. However, at the 6-month postoperative time point, the healthy and overweight groups show significant differences in the JOA scores from the obese group.

### Gait analysis

Figs 3 and 4 show the gait analyses preoperatively, 6 months postoperatively, and 1 year postoperatively. Preoperatively, no significant differences were observed between the healthy, overweight, and obese groups. Regarding walking speed, the obese group had a lower walking speed compared with the healthy and overweight groups, at 6 months postoperatively as well as 1 year postoperatively. In terms of stride length, the obese group had a lower stride length compared with the healthy and overweight groups at 6 months and 1 year postoperatively.

**Fig 3.**
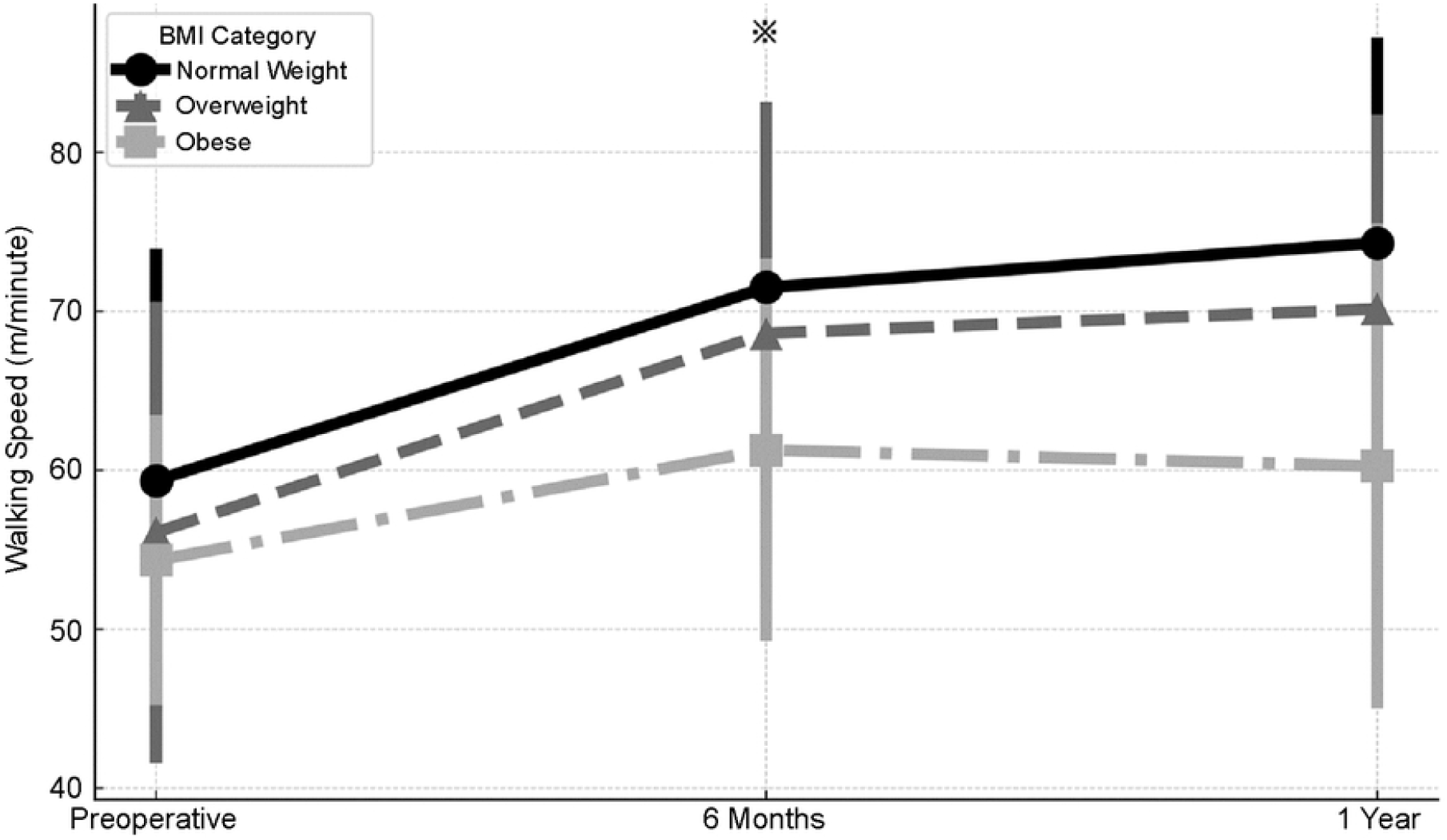
Walking speed. Preoperatively, no significant differences are observed among the healthy, overweight, and obese groups. At 6 months postoperatively, the obese group has a slower walking speed, compared with the healthy group, and at 1 year postoperatively, the obese group has a slower walking speed, compared with both the healthy and overweight groups (*p* < 0.05).

**Fig 4.**
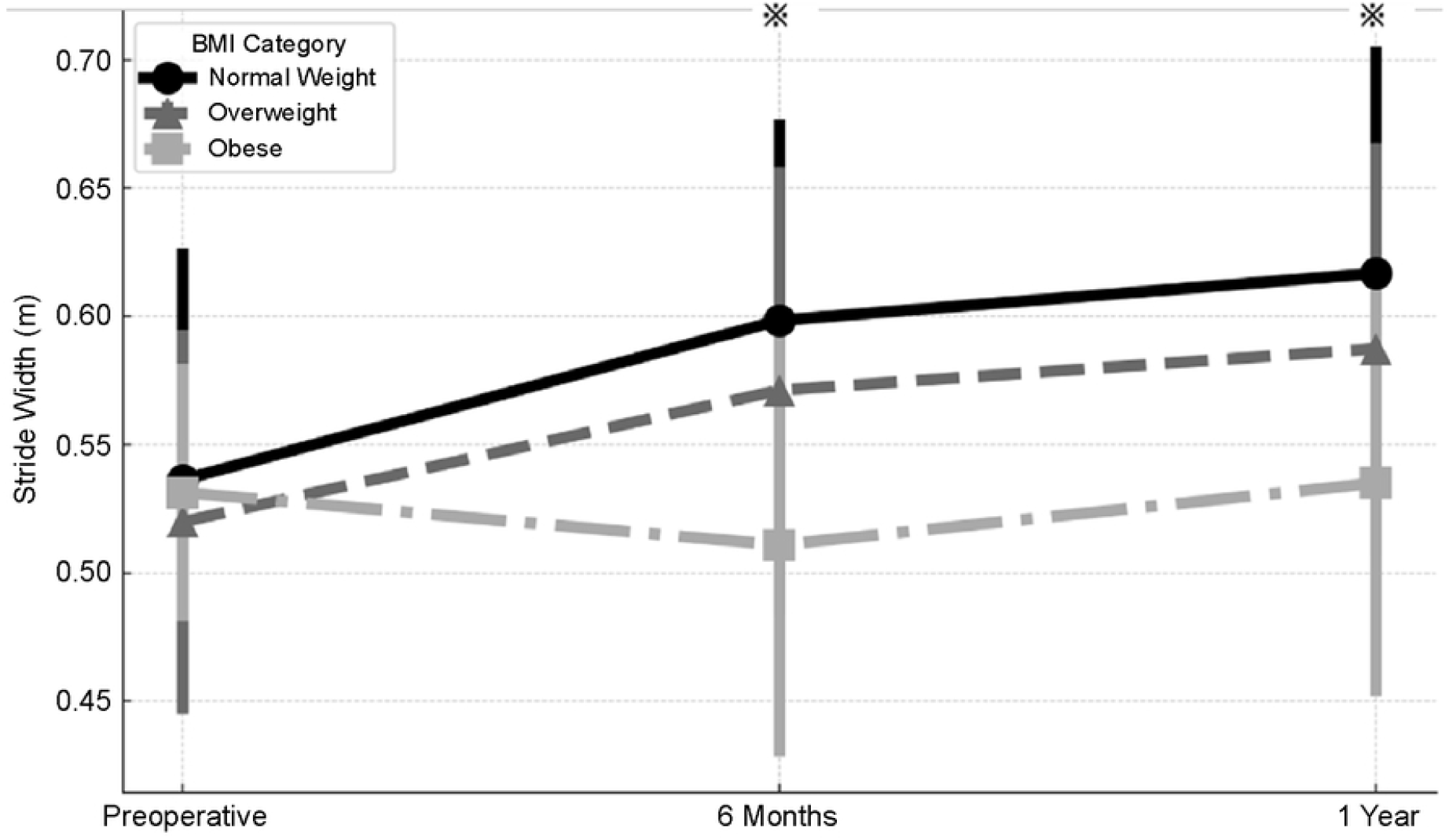
Slide width. Preoperatively, no significant differences in stride length are observed among the healthy, overweight, and obese groups. Six months and 1 year postoperatively, the obese group has shorter stride length, compared with the healthy and overweight groups.

## Discussion

This study’s results showed no significant differences in the JOA hip scores among the groups at 1 year post-surgery. However, gait analysis indicated significantly lower performance in the obese group at 6 months and 1 year after surgery. Recent trends toward shorter hospital stays for THA and the adoption of outpatient surgery in countries, such as the United States [19], aims to reduce healthcare costs. Studies have shown no significant differences in outcomes between inpatient and outpatient hip arthroplasty [20].

### Functional recovery and outcome evaluation in patients with obesity after THA

Although an increase in BMI was associated with complications and delayed recovery of postoperative walking function, the JOA hip scores at 1 year post-surgery show no significant differences among the BMI groups. This suggests that patients with obesity can achieve a functional recovery than those without obesity over the long term [21]. This outcome indicates that surgical intervention significantly enhances the quality of life of patients by effectively reducing pain and improving their range of motion. Despite early challenges, such as increased exercise load and delays in rehabilitation [21–23], which are exacerbated by obesity, the postoperative improvements in subjective symptoms such as pain and mobility post-surgery are significant. Although we did not assess pain using a separate visual analog scale (VAS), the JOA hip score includes a pain component, which partially reflects postoperative pain levels. However, future studies should incorporate more detailed pain assessments, such as gait-related VAS, to better quantify the relationship between pain and gait function.

The JOA score, which reflects patients’ pain and daily life inconvenience [24], highlights that enhancing the quality of daily living is a critical outcome for patients, distinct from the pace of objective functional recovery. These findings emphasize the complexity of the impact of obesity on postsurgical recovery and the need to incorporate these considerations into rehabilitation planning to optimize recovery trajectories.

### Challenges and strategies in rehabilitation for patients with obesity after THA

Gait analysis shows that the obese group had inferior walking ability than did the other BMI groups at both 6 months and 1 year postoperatively. This indicates that obesity significantly impedes walking function recovery [12,15]. This reduction in walking speed and stride length severely impacts activity levels and daily life independence, thereby highlighting the need for tailored rehabilitation interventions for patients with obesity. During the short-term recovery phase, focused rehabilitation strategies, including pain management, joint protection, and muscle strength maintenance, are essential for addressing delays and increased discomfort during exercise [25,26].

Over the long term, continued deficits in walking ability require a sustained focus on enhancing walking efficiency and exercise tolerance. This can be achieved through gradual increases in exercise load, posture adjustments, and balance training. Additionally, integrating weight management and nutritional counseling is essential for alleviating joint stress and optimizing rehabilitation outcomes. This study highlights the importance of personalized rehabilitation programs that consider each patient’s unique physical condition, comorbidities, living environment, and personal goals, to effectively address the challenges faced by patients with obesity during post-THA rehabilitation.

### Limitations

This study provides several valuable insights into the impact of BMI on functional recovery after THA for OA. However, some limitations must be considered. First, the limited sample size warrants caution in generalizing the findings. Additionally, the influence of specific demographic characteristics may skew the results, thereby raising questions about their applicability to other populations. Furthermore, the 1-year postoperative observation period may not fully capture the long-term effects of THA.

The lack of standardization in the execution of surgery and rehabilitation programs can introduce variability in study outcomes. Additionally, while the importance of using both subjective and objective assessments is recognized, the limitations of these assessment methods and the potential for bias in subjective evaluations must be considered. Furthermore, the impact of external factors, such as patients’ lifestyle habits and socioeconomic status, which may have affected the interpretation of the findings on the results, has not been fully evaluated. Additionally, although the JOA hip score includes a pain subdomain, the lack of a specific gait-related pain evaluation, such as a VAS score during walking, is a limitation. Furthermore, while adjacent joint OA was excluded radiographically, subtle biomechanical influences from unmeasured joint or spinal conditions cannot be entirely ruled out.

## Conclusion

This prospective study assessed the impact of BMI on functional recovery following THA for OA. Patients with obesity experienced significant delays in recovery, particularly in walking speed and stride length, during the first postoperative year. Despite these delays, the JOA hip scores showed no significant differences among the BMI groups at 1 year postoperatively, suggesting that clinical outcomes are comparable across groups over time.

These findings highlight the need for personalized rehabilitation strategies for patients with obesity to address specific challenges and optimize the long-term success of THA.

## Data Availability

All relevant data are within the manuscript.

## Acknowledgements

None.

